# Artificial intelligence in clinical genetics: current practice and attitudes among the clinical genetics workforce

**DOI:** 10.1101/2025.04.30.25326673

**Authors:** Amanda M Berkstresser, Suzanna E. Ledgister Hanchard, Daniela Iacaboni, Kevin McMilian, Dat Duong, Benjamin D. Solomon, Rebekah L. Waikel

**Author notes:** Correspondence: Rebekah L. Waikel, PhD, CGC, *National Human Genome Research Institute,* Building 10 - CRC, Suite 3-2551, 10 Center Drive, Bethesda, MD 20892.

## Abstract

**Purpose:** Artificial intelligence (AI) applications for clinical genetics hold the potential to improve patient care through supporting diagnostics and management as well as automating administrative tasks, thus enhancing and potentially enabling clinician/patient interactions. While the introduction of AI into clinical genetics is increasing, there remain unclear questions about risks and benefits, and the readiness of the workforce.

**Methods:** To assess the current clinical genetics workforce’s use, knowledge, and attitudes toward available medical AI applications, we conducted a survey involving 215 US-based genetics clinicians and trainees.

**Results:** Over half (51.2%) of participants report little to no knowledge of AI in clinical genetics and 64.3% reported no formal training in AI applications. Formal training directly correlated with self-reported knowledge of AI in clinical genetics, with 69.3% of respondents with formal training reporting intermediate to extensive knowledge of AI vs. 37.5% without formal training. Most participants reported that they lacked sufficient knowledge of clinical AI (83.4%) and agreed that there should be more education in this area (97.6%) and would take a course if offered (89.3%). The majority (51.6%) of clinician participants said they never used AI applications in the clinic. However, after a tutorial describing clinical AI applications, 75.8% reported some use of AI applications in the clinic. When asked specifically about clinical AI application usage, the majority of clinician participants used facial diagnostic applications (54.9%) and AI-generated genomic testing results (62.1%), whereas other applications such as chatbots, large language models (LLMs), pedigree or medical summary generators, and risk assessment were only used by a fraction of the clinicians, ranging from 11.1 to 12.5%. Nearly all participants (94.6%) reported clinical genetics professionals as being overburdened.

**Conclusion:** Further clinician education is both desired and needed to optimally utilize clinical AI applications with the potential to enhance patient care and alleviate the current strain on genetics clinics.

## Introduction

The use of AI in healthcare is increasing, such as via AI-generated interpretations of pathology slides and radiology image^1^, AI-based scribes for documentation, and applications for protein modeling to support therapeutic research^2^. Educational uses in healthcare include virtual simulation and training for surgical and other procedures, teaching clinicians to identify imaging findings such as hip fractures in plain X-rays or melanomas in histopathology, and natural language processing (NLP) to support trainee evaluation of geriatric patients ^3^. In genetics education, a recent study explored the use of generative AI to make realistic images to train pediatric residents to recognize children with two genetic conditions^4^. In clinical genetics settings, examples of uses of AI include clinical chatbots like GIA (Genetic Information Assistant) to facilitate genetic information gathering for patients^5^, or to provide genetic breast cancer education to patients^6^, web-based applications like Face2Gene (https://www.face2gene.com/) to help with differential diagnosis generation, and the use of AI-assisted search engines and large language models (LLMS) for both patients and clinicians to look up pertinent clinical information^7^, AI-based risk assessment to calculate disease risk for diseases such as cancer^8–10^ and cardiovascular disease^11^, AI generated medical summaries and pedigree^8,12,13^, and AI-supported diagnoses from sequence data^14,15^.

In the field of clinical genetics, AI has the potential to help address intrinsic challenges, including a shortage of medical geneticists and genetics professionals relative to patient need^16^, the lack of standardized or sufficiently comprehensive genetics training in medical schools, residencies and beyond, and the rapidly evolving clinical landscape, where new disease-causing genes and genetic conditions are constantly being described and expanded, new management guidelines are frequently disseminated, and new treatments are emerging^2^. Despite the potential benefits of AI in clinical genetics, uses appear to be highly variable^17^. This variability may be multifactorial, including related to disparate views, knowledge, and experience with AI, as well as different rules and regulations regarding AI’s uses such as in different geographic regions and medical or individual hospital systems.

To date, there are no published studies specifically addressing uses and perception of AI amongst genetics clinicians, though there are relatively recent survey studies looking broadly at AI and healthcare. For example, a survey of 1600 researchers showed that the majority view AI as being potentially beneficial to improve productivity, but noted concerns such as overreliance on AI systems and fears about bias or discrimination, as well as the potential for fraud or irreproducible research^18^. The views of patients also demonstrate mixed responses about the integration of AI in healthcare; patients preferred AI-generated messaging responses compared to those written by physicians^19^, but reported concerns about issues such as privacy, data integrity, and autonomy^20^. There have been some healthcare provider studies, with many focused on the field of radiology, a specialty that has become increasingly dependent on AI for diagnostic purposes. Surveys of radiologists demonstrate support for the use of AI, but respondents reported insufficient education and resources to optimally integrate AI into clinical practice^21,22^. Similar views were described by general healthcare providers, who were generally positive about AI but who also cited the need for more education and practical experience^23–25^.

In clinical genetics, AI holds the potential to accelerate and improve diagnostics, as well as to free clinicians from time-consuming administrative tasks, such as involving documentation and insurance-related paperwork. However, there are important questions regarding the use of AI in the field. To explore the uptake and attitudes of clinical genetics professionals, we surveyed medical geneticists, genetic counselors, nurse practitioners, physician assistants, nurses, genetics residents/fellows, and genetic counseling students regarding their utilization of AI in clinical genetics practice, including exploring awareness of their current use of AI (e.g., whether they were aware that certain commonly used applications involve AI), as well as their attitudes, concerns, and perceptions about this type of technology.

## Materials and Methods

### Survey instrument

An online survey was designed in Qualtrics (Provo, Utah, United States); the survey included the following types of questions: demographic, clinical AI education and experience, and AI utilization perception. Some questions used a 4-point Likert scale (strongly disagree, disagree, agree, strongly agree). The survey also included a tutorial describing current popular applications of AI in genetics clinics (selected based on discussions with a variety of genetics professionals at different career stages), including facial diagnostics, genomic testing, LLM - based chatbots, clinical chatbots, pedigree and medical summaries, and risk assessment. The same four clinical AI perception questions (self-use, colleague use, accessibility, and beneficiality of AI applications) were asked both before and after the tutorials to test whether participants’ perceptions would change after being presented with additional information about AI applications in the clinic. Survey participants could proceed through the survey without needing to respond to each question before moving to the next question. Therefore, the reported number of respondents varies per question. The survey can be found at: https://github.com/rlwaikel/AI_Clinical_Genetics.

### Participant recruitment

Board-certified/board-eligible clinical genetics professionals (MD, DO, GC, RN, NP, or PA) and genetics trainees (student, resident, or fellow) were recruited directly via email or indirectly via a flyer disseminated to training program directors and professional organizations that provided a link to a survey form. Contact information for potential participants was obtained via professional networks and online sources. Survey data were collected from October 2023 to June 2024. Of the 227 individuals who opened the survey, 215 progressed beyond demographic questions and provided responses to clinical AI questions. Only surveys from individuals who completed at least the first section, which included clinical AI education and experience, were included in the analysis.

The study was approved by the Bay Path University IRB (IRB# 2023_Berkstresser).

### Data analysis

All survey data were collected through Qualtrics and stored in Excel. No personal information was collected or saved. For analyses, participants were divided into two main groups: clinicians (including medical genetics residents and fellows) and genetic counseling students. Likert scale results were binned into two categories: agree (strongly agree and agree responses) and disagree (disagree and strongly disagree responses). We applied 2-sample t-test to compare perception responses for both before/after educational intervention and clinicians versus genetic counseling students. To determine the correlation of self-reported knowledge and training, we summed the “extensive” and “intermediate” response as one single answer choice while keeping “little to no knowledge” as its own choice; allowing us to compare responses for what we considered as high versus low knowledge. Then, we divided the participants based on whether they received any training (clinical program and/or workshop/seminar). Logistic regression was done for each participant group with self-reported training as covariate and self-reported knowledge as outcome.

## Results

### Participant demographics

Participants represented individuals from 36 different states and Washington, D.C. (Figure 1a). Participants, all of whom worked primarily in genetics, included 148 board-eligible or board-certified professionals (83 genetic counselors, 54 medical geneticists, 9 physician assistants, 1 nurse practitioner, and 1 registered nurse), 6 medical genetics residents or fellows, and 61 genetic counseling students (Figure 1b). Most participants reported primarily clinical or a combination of clinical and research roles (68.2%;101/148) with primarily research (14.9%; 22/148) and primarily education (13/148) being the next most common position types (Figure 1c). Most non-trainee professional respondents reported their primary place of employment as an academic medical or research center (73.6%; 109/148) (Figure 1d). Additional demographic information (e.g., related to age and experience) is provided in Supplemental Figure 1.

**Figure 1.**
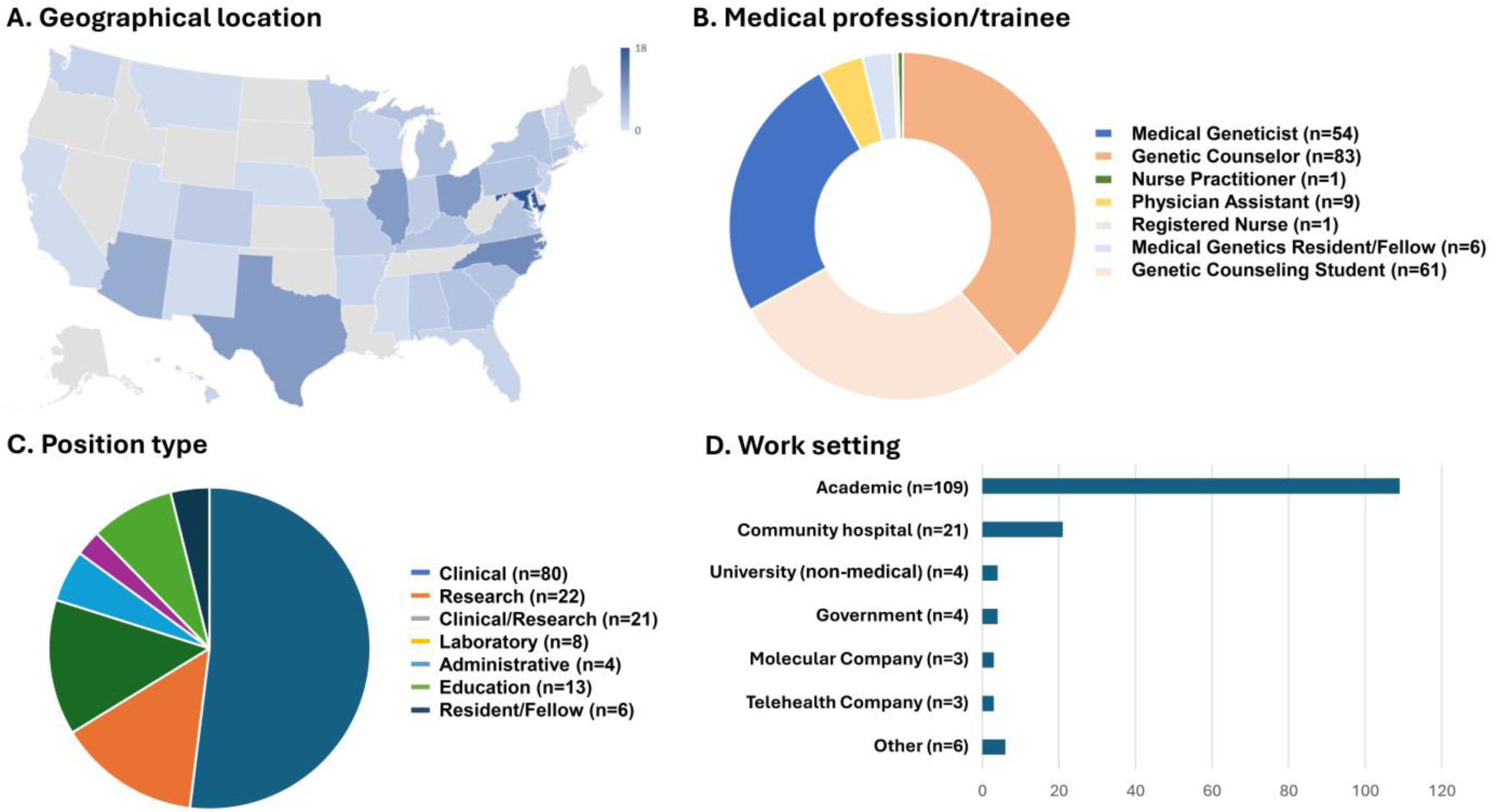
Survey participant demographics. Participants broadly represent 36 states and Washington, DC (A). The largest three cohorts of participants were medical geneticists (54), genetic counselors (83), and genetic counseling students (61) (B). Most professional participants described their work as primarily clinical (80) (C), and most worked in academic medical or research centers (D).

### AI education and self-reported knowledge

Approximately one-third of respondents (35.5%; 75/211) reported receiving some training in AI; most of these (31.8%; 67/211) reported that they received this training through a conference or workshop, with less (8.1%; 19/211) receiving training as part of their professional program curriculum; 5.2% (11/211) of participants reported training in both (Figure 2). More than half of respondents (51.2%; 108/211) described themselves as having little to no knowledge regarding the application of AI in clinical genetics. Relatively few (1.9%; 4/211) rated themselves as having extensive knowledge of AI in clinical genetics and 46.9% (99/211) rated their knowledge as intermediate. Those who received either type of AI training were more likely to rate themselves as having intermediate or extensive knowledge (69.3%; 52/75) versus those without training (37.5%; 51/136), p=6.5x10^-6^ (Figure 2).

**Figure 2.**
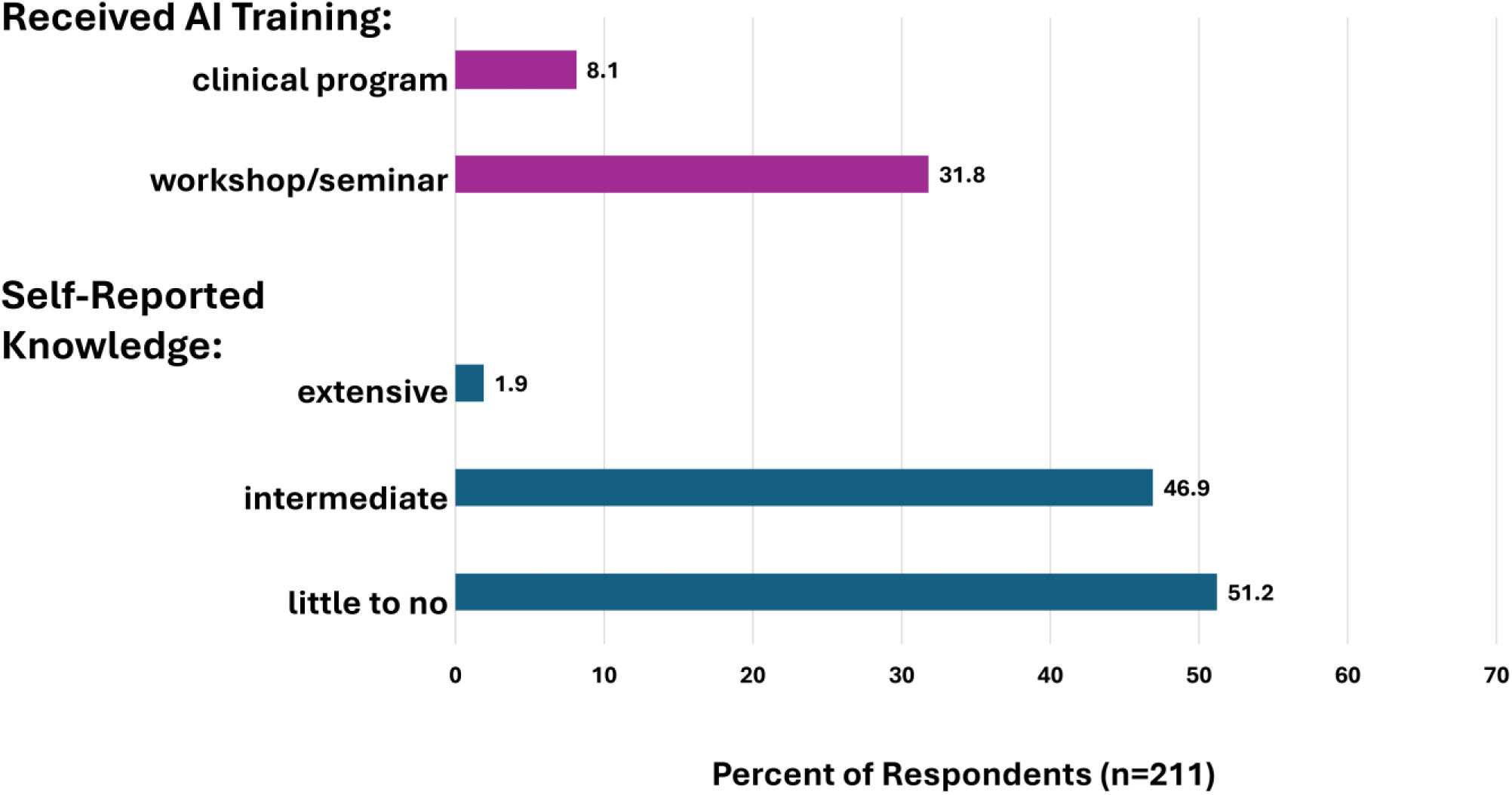
Clinical genetics AI training and self-reported knowledge. Only 8.1% received training as part of their clinical program and 31.9% received training in a workshop or conference, collectively representing 35.7% of participants. Just over half of participants reported their knowledge level to be little to none. There was a positive correlation between having received training and intermediate/advanced self-reported knowledge, p=6.5x10^-6^.

### Applications for AI in clinical genetics

To query whether clinician participants were aware of applications of AI in clinical practice, we provided an informational tutorial on several current uses including applications involving facial diagnostics, genomic testing, LLM-based chatbots, clinical chatbots, pedigree and medical summaries, and risk assessment. We assessed awareness of AI uses both directly (self-reported; described here) and indirectly (described in the next section). A paragraph describing each application was provided, followed by 2 questions with binary answers (yes/no), which asked whether the clinician had ever used this type of AI application and whether the clinician was aware that the application was AI-based. Clinician participants reported the greatest use of AI in facial diagnostics (54.9%; 84/153; Supplemental Figure 2a) and genomic testing (62.1%; 96/153; Supplemental Figure 2b). Usage of the other four applications ranged from 11.1-12.5% of respondents (Supplemental Figure 2c-f). The majority of clinicians were aware that each of the six example applications were AI-based, with some variability, ranging from the least known (risk assessment) (52.3%; 80/153; Supplemental Figure 2f) to the best known (LLMs) (96.1%; 147/153; Supplemental 2d).

Additionally, we also asked genetic counseling students about their knowledge and anticipated use of these applications in their future practices. Genetic counseling students reported greater anticipated use of clinical chatbots, LLMs, AI prepared notes, and risk assessment applications than current clinicians reported current use (Supplemental Figure 2c, d, e, f).

### Frequency of use, accessibility, and beneficiality of AI applications in the genetics clinic before and after clinical AI applications tutorial

To indirectly assess clinicians’ awareness of the use of clinical AI applications, we determined the change in responses of a series of four questions before and after exposure to the AI application tutorial described above. These questions were broadly about AI in the clinic (not specific examples as described above) and included asking how often the participant engages with AI in general in their clinic, as well as how often the participant perceives that colleagues engage with AI in general. Prior to the educational content, more than half of clinician participants (51.6%; 77/149) reported never using AI in the clinic, but 93% (139/149) perceive their colleagues using it to some degree, with the most common response being “rarely use” (Figure 3). After exposure to examples of AI applications in the clinic, levels of self- and perceived colleague use increased to 75.8% (113/149) reporting some level of self-use and 96.6% (144/149) perceived colleague use with the most common response as weekly use. P-values for self- and perceived colleague-use were 5.2x10^-10^ and 1.5x10^-5^ compared to the reports of use prior to the educational exposure, respectively.

**Figure 3.**
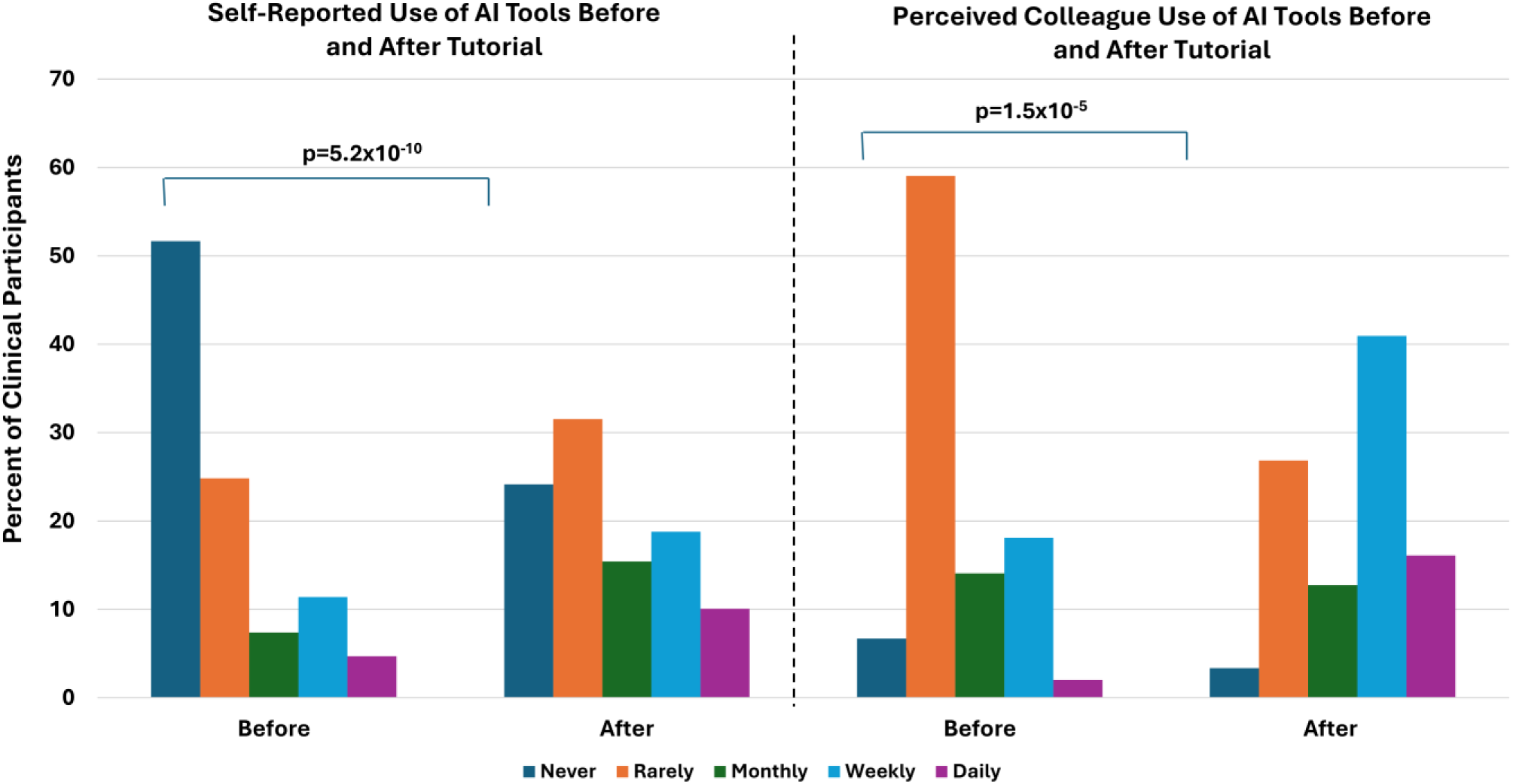
Frequency of AI tool use in the genetics clinic before and after tutorial (n=149). The majority of clinician participants reported never using AI tools in the clinic, whereas after a tutorial describing several uses of AI tools in the genetics clinic, there was a significant reduction in those reporting never using AI tools. Clinician participants also perceived that their colleagues use AI tools more than they do. As with self-use of AI tools, after the tutorial, participants reported more frequent use of AI tools amongst their colleagues.

Both prior to and after the tutorial, we asked broadly whether clinical AI applications are accessible and whether these applications are beneficial. One-third of participants (33.3%; 50/150) responded that clinical AI applications were not accessible to them (Figure 4a). After being provided information and explanations about clinical AI applications, only 8% (12/150) reported that AI applications were not available (p=1.6x10^-10^). Participants were also asked to provide their opinion on how beneficial they perceived clinical AI applications to be. Most clinician participants (86.5%; 123/141) reported AI applications to be beneficial or highly beneficial (Figure 4b). After exposure to educational information, participants reported a significantly more favorable view, with 97.2% (137/141) reported beneficial or highly beneficial (p=7.4x10^-5^). Additionally, the majority of participants who have used clinical AI (65.1%; 121/186) reported that AI applications were somewhat easy to very easy to use.

**Figure 4.**
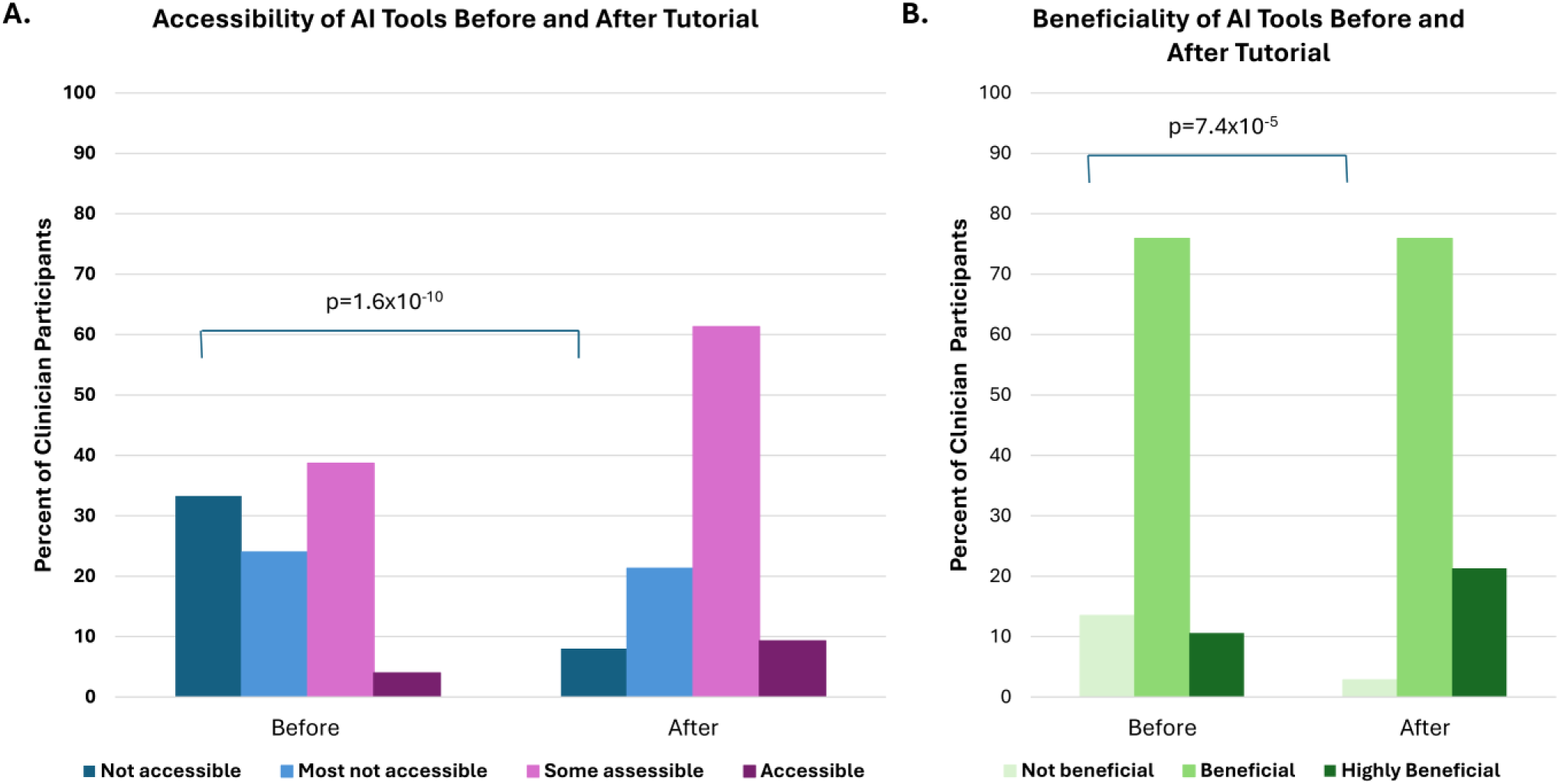
Perception of accessibility and beneficiality of AI tools in the genetics clinic before and after tutorial. Prior to the tutorial describing examples of uses of AI tools in the genetics clinic, most participants responded that AI tools were either not accessible or that most were not accessible. After the tutorial, most clinician participants reported that at least some tools were accessible (A; n=150). Before the tutorial, 13.5% reported AI tools as not beneficial, whereas after tutorial less than 3% reported AI tools as not beneficial (B; n=141).

### Comfort with the use of AI in clinical genetics

Analysis of responses showed that more than half of clinician participants indicated that they are comfortable using AI as part of their clinical practice (62.3%; 129/207), and felt that their patients would be comfortable using AI (57.1%; 116/203), as signified by an agree or strongly agree responses (Table 1). When asked specifically about types of use, clinicians responded that were more comfortable with patients obtaining general genetics information (65.4%; 134/205) as compared to their patients using AI to undergo pretest genetic counseling (41.9%; 86/205) (Table 1).

**Table 1.** Likert scale questions assessing the clinical genetics workforce’s opinions and attitudes toward.

| Likert Scale Questions | Agree/<br>Strongly<br>Agree | Disagree/<br>Strong<br>Disagree |
| --- | --- | --- |
| **AI Knowledge and Education** |  |  |
| I currently <u>lack sufficient knowledge</u> about AI in medical genetics. n=205 (%) | 171 (83.4) | 34 (16.6) |
| There <u>should be further curriculum</u> and education about the use of AI in clinical care. n=206 (%) | 201 (97.6) | 5 (2.4) |
| If offered a <u>class about AI</u> in clinical genetics, I would take it. n=206 (%) | 184 (89.3) | 22 (10.7) |
| **Comfort with AI in Clinical Practice** |  |  |
| I am <u>comfortable</u> using AI in my clinical genetics practice. n=207 (%) | 129 (62.3) | 78 (37.7) |
| My <u>patients</u> would be <u>comfortable with the use of AI</u> in clinical genetics. n=203 (%) | 116 (57.1) | 87 (42.9) |
| I am <u>comfortable</u> with my patients <u>using AI to obtain general genetics information</u>. n=205 (%) | 134 (65.4) | 71 (34.6) |
| I am <u>comfortable</u> with my patients using <u>AI to undergo pretest genetic counseling</u>. n=205 (%) | 86 (41.9) | 119 (58.1) |
| **AI Safety and Reliability** |  |  |
| AI provides reliable outputs. n=203 (%) | 144 (70.9) | 59 (29.1) |
| I am concerned AI will <u>not be used responsibly</u> in clinical genetics. n=206 (%) | 102 (49.5) | 104 (50.5) |
| There needs to be <u>further regulations</u> for AI before I incorporate it into my genetics practice. n=205 (%) | 168 (81.9) | 37 (18.1) |
| AI can be safely used in the handling of confidential data. n=204 (%) | 130 (63.7) | 74 (36.3) |
| **Clinical Genetics Clinic Workload** |  |  |
| Clinical genetics professionals are over-burdened. n=204 (%) | 193 (94.6) | 11 (5.4) |
| I am willing to delegate work to other people. n=202 (%) | 189 (93.6) | 13 (6.4) |
| I am willing to delegate work to AI. n=201 (%) | 162 (80.6) | 39 (19.4) |
| AI is an appropriate solution to relieving workload. n=202 (%) | 147 (72.8) | 55 (27.2) |
| **Attitudes Toward AI Applications** |  |  |
| I have felt forced to utilize AI in my practice. n=207 (%) | 15 (7.2) | 192 (92.8) |
| Using AI as an aid is outside my scope of training/practice. n=206 (%) | 57 (27.7) | 149 (72.3) |
| <u>AI is not ready</u> to be used in clinical genetics. n=204 (%) | 52 (25.5) | 152 (74.5) |
| AI is inappropriate for use as a diagnostic support application. n=208 (%) | 33 (15.9) | 175 (84.1) |
| The development of new medical genetics AI applications requires input from clinical genetics professionals. n=206 (%) | 205 (99.5) | 1 (0.5) |

### AI reliability and safety

We asked a series of questions to better understand perception of reliability, responsible use, regulations on use, and ability to handle confidential (private, sensitive) data securely (Table 1). Most participants (70.9%; 144/203) agreed or strongly agreed that AI provides reliable outputs. Just over half of clinician respondents (50.5%; 104/206) were not concerned that AI would be used irresponsibly in clinical genetics. The majority (81.9%; 168/205) also reported that there needs to be further regulations for AI before incorporating it into their clinical practices. A majority (63.7%; 130/204) agreed that AI can safely handle confidential data.

### Genetics clinic workload

Most participants (94.6%; 193/204) agreed or strongly agreed with the statement that clinical genetics professionals are overburdened (Table 1). Related to possible ways to address this issue, most participants indicated that they were willing to delegate work to other people (93.6%; 189/202) and to AI (80.6%; 162/201). When asked if AI is an appropriate solution to relieving workload, a similar number of participants agreed or strongly agreed with that statement (72.8%; 147/202) as indicated that they were willing to delegate work to AI (r=0.71).

### General attitudes toward AI

To better understand general attitudes towards AI in clinical genetics, we asked a series of negatively stated questions (Table 1). When asked if the participant felt forced to utilize AI, the majority (92.8%; 192/207) either disagreed or strongly disagreed. When presented with the statement “Using AI as an aid is outside of my scope of training/practice”, the majority (72.3%; 149/206) disagreed or strongly disagreed. The majority of participants (74.5%; 152/204) also disagreed with the statement that “AI is not ready to be used in clinical genetics.” Additionally, the majority (84.1%;175/208) disagreed or strongly disagreed with the statement “AI is inappropriate for diagnostic purposes.” Nearly all participants (99.5%; 204/206) agree that the development of new medical genetics AI applications requires input from clinical genetics professionals.

### AI knowledge and education

A major concern reported by participants is that they lack sufficient knowledge about AI in medical genetics; 83.4% (171/205) agreed or strongly agreed with this statement (Table 1).

Most participants (97.6%; 201/206) agreed or strongly agreed that there should be further curricular and educational opportunities about the use of AI in clinical care. Most (89.3%; 184/206) also agreed or strongly agreed that they would take a class about AI in clinical genetics, if offered.

### Response differences between genetic counseling students and practicing clinicians

There are a few notable differences between genetic counseling students and practicing clinicians, with students reporting greater concerns regarding patient use, responsible use, and regulations of AI, while being more open to delegating work to AI (Supplemental Table 1).

Genetic counseling students were similarly comfortable with self-use of AI in their genetics practice as compared to current clinicians. However, their comfort with their patients’ use of AI for clinical genetics purposes was significantly lower when compared to clinicians (33.9%; 19/56 vs. 66%; 97/147, p=4.4x10^-5^). Specifically, lower comfort levels were observed in students as compared to clinicians for general genetics information (48.2%; 27/56 vs. 71.8%; 107/149, p=2.8x10^-3^) and pretest counseling (23.2%; 13/56 vs. 49%; 73/149, p=3.7x10^-4^) (Supplemental Table 1).

Like clinicians, the majority of genetic counseling students agreed that AI provides reliable outputs and can be safely used in handling data that should be kept confidential. However, genetic counseling students were significantly more concerned with responsible use and regulations of AI in the genetics clinic than clinicians (63.2%; 35/57 vs 44.3% 66/149, p=9.3x10^-3^) and (96.5%; 55/57 vs 76.4%; 113/148, p=4.8x10^-6^), respectively (Supplemental Table 1).

While students and clinicians had similar responses about the appropriateness of AI as a workload solution, genetic counseling students were more willing than clinicians to delegate work to AI (92.7%; 51/55 vs. 76%; 111/146, p= 1.1x10^-3^) (Supplemental Table 1).

### Response differences among clinical professions

The largest two participant cohorts of clinical professionals were genetic counselors (n=83) and medical geneticists (n=55). Other groups with more than one participant were physician assistants (n=9) and medical genetics residents and fellows (n=6). The small cohort size of these latter two groups made statistical comparison difficult, and therefore only trends will be mentioned.

These two larger cohorts tended to agree on most responses (Supplemental Tables 2 and 3) except for accessibility of AI applications and four Likert scale questions (Supplemental Table 4). With respect to perceived accessibility, a greater percentage of medical geneticists found these applications accessible when compared to genetic counselors (57.4; 31/54 vs 32.1; 27/84; p=0.005). Medical genetics residents and fellows also found the application to be more accessible, with 66.7% (4/6) reporting applications as accessible, whereas physician assistants reported similar accessibility as genetic counselors (33.3%; 3/9) (Supplemental Table 2).

Genetic counselors were more comfortable with patients using AI to obtain general genetics information than medical geneticists (81%; 64/79 vs 64.2%; 34/53; p=0.03), as well as physician assistants and residents/fellows (55.6%; 5/9 and 50%; 3/6). Significantly more genetic counselors agreed that AI provides reliable outputs than medical geneticists (84.6%; 66/78 vs. 51.9%; 27/52; p=3.42x10^-5^). Both physician assistants and resident/fellows also reported more agreement with the statement regarding reliability (62.5%; 5/8 and 83.3%; 5/6). A greater percentage of genetic counselors than medical geneticists agreed that further regulation on clinical AI is needed (97.5%; 77/79 vs 67.9%; 36/53; p=2.05x10^-8^). Physician assistants unanimously agreed about the need for regulations (100%: 9/9), whereas residents/fellow had a similar agreement rate as medical geneticists (66.7%; 4/6). Genetic counselors were also more likely to agree to with the statement “I currently lack sufficient knowledge about AI in medical genetics” compared to medical geneticists (85.9%; 67/78 vs 67.9%; 36/53; p=0.01). Agreement rates for residents/fellows were similar to those of genetic counselors (83.3%; 5/6), whereas the agreement was lower with physician assistants (66.7%; 6/9).

## Discussion

Despite reporting that AI applications are beneficial to clinical genetics, the average participant reported rarely to never using AI applications in their clinical practice. Interestingly, participants perceived that their colleagues have greater use than themselves. There are several clues in the survey results that point to possible reasons limiting AI use, including accessibility of AI applications, education, and hesitancy to use AI for all aspects of clinical genetics, including return of uncertain and positive genetic testing results. Participants acknowledged that the use of AI is part of clinical genetic practice and that it was reliable and safe. Additionally, participants indicated that AI has the potential to relieve workloads and reported willingness to delegate work to AI applications.

A novel aspect of our survey was the inclusion of tutorials describing current AI tools (facial diagnostics, genomic testing, LLM -based chatbots, clinical chatbots, pedigree and medical summaries, and risk assessment) that may be used in genetics clinics. This facilitated both the goal of enhancing participants’ knowledge of AI uses in clinical genetics and also provided a way to assess awareness of AI usage by analyzing differences in answers to the same four questions (self-use, colleague use, accessibility, and beneficiality of AI tools) both before and after exposure to the tutorial. Interestingly, after the tutorial, participants reported a significant increase in self-use, perceived colleague use, accessibility, and beneficiality, suggesting that the participants were not fully aware of AI in their clinical practice prior to the survey.

There were some notable differences between participant groups. Genetic counseling students demonstrated more permissive views about self-use of AI in the clinic, such as a greater willingness to delegate work to AI and higher anticipated AI application use rate than clinicians but did have unique concerns. They were cautious about their patients’ use of AI for all aspects of genetic counseling including general information gathering and pretest counseling, as well as return of results, which were of greater concern for all groups. Students were also more concerned about responsible use and regulations of AI. Like students, practicing genetic counselors were also more concerned about the need for regulations of clinical AI as compared to the larger participant pool, but did not echo the other concerns mentioned by genetic counseling students. When compared to their medical geneticist colleagues, genetic counselors had greater comfort with patients using AI applications, which might be attributed to a greater confidence in the reliability of AI outputs.

To date, there is no standard AI curriculum as part of medical genetics training programs (https://www.acgme.org/programs-and-institutions/programs/common-program-requirements/). While only a small fraction of our participants received AI education in their clinical training programs, a greater number sought out training through a conference or workshop. Not surprisingly, those receiving training were more likely to report intermediate or advanced knowledge of clinical AI. Most participants indicated a need for more AI education, with nearly 90% of participants indicating they would take a class on clinical AI.

### Limitations

This study was dependent on self-reporting, which is inherently subjective and may be skewed by variances in self-perceptions and interpretations of prompts. Similarly, some items were asked broadly, which could allow misinterpretation of the intention. An example would be the opinion that “AI is not ready to be used in clinical genetics.” This item used AI as a general term, allowing participants to potentially apply different interpretations when selecting their response. This study also offers a brief “snapshot in time” of a limited number of participants. As AI is being integrated into many parts of healthcare and other parts of society and is receiving considerable attention in the lay and scientific press, it is likely that attitudes and uses about AI will continue to evolve.

## Conclusion

Our research sought to answer questions regarding the current utility of AI in clinical genetics and clinicians’ views about its utilization. While survey data indicated that genetics professionals overall view AI as beneficial, there were still hesitancies, specifically regarding the implementation of LLMs such as ChatGPT, Gemini, or Claude, or the use of AI for particularly critical conversations such as pretest genetic counseling. Still, responses indicated AI as a possible solution to combating workload demands, and participants were willing to learn more if given the opportunity. The many potential applications of AI are continuing to evolve as this technology becomes increasingly integrated into healthcare. Genetics professionals must stay up to date in a constantly changing field, and as indicated by respondents, AI is not outside the scope of practice. Further exploration of regulations and the establishment of professional guidelines is needed to ensure the powerful technology is being used both efficiently and ethically; this process will ideally be guided in cooperation with the larger genetics ecosystem.

## Availability of data and materials

The survey can be found at: https://github.com/rlwaikel/AI_Clinical_Genetics. Raw survey data are available upon written request.

## Acknowledgements

This research was supported by the Intramural Research Program of the National Human Genome Research Institute of the National Institutes of Health and Bay Path University Genetic Counseling Program, and used the computational resources of the National Institutes of Health High-Performance Computing Services Biowulf cluster.

## Contributions

Concept and design: Berkstresser, Waikel, Solomon, Duong, Iacomboni, and McMilian

Acquisition, analysis, or interpretation of data: All authors

Drafting of the manuscript: Berkstresser, Ledgister Hanchard, Waikel, Solomon

Statistical analysis: Berkstresser, Waikel, Duong

Obtained funding: Solomon

Administrative, technical, or material support: Iacomboni, McMilian, Duong

Supervision: Waikel, Solomon, and Iacomboni

Conflict of Interest Disclosures: BDS is the co-Editor-in-Chief of the American Journal of Medical Genetics and receives textbook royalties from Wiley publishing. Both editing/publishing activities are conducted as an approved outside activity, separate from his US Government role. No other authors have conflicts of interest or additional acknowledgements. RLW had full access to all the data in the study and takes responsibility for the integrity of the data and the accuracy of the data analysis.

