## Supplemental Figures 1-3 for "Artificial intelligence in clinical genetics: current practice and attitudes among the clinical genetics workforce"

### Slide 1
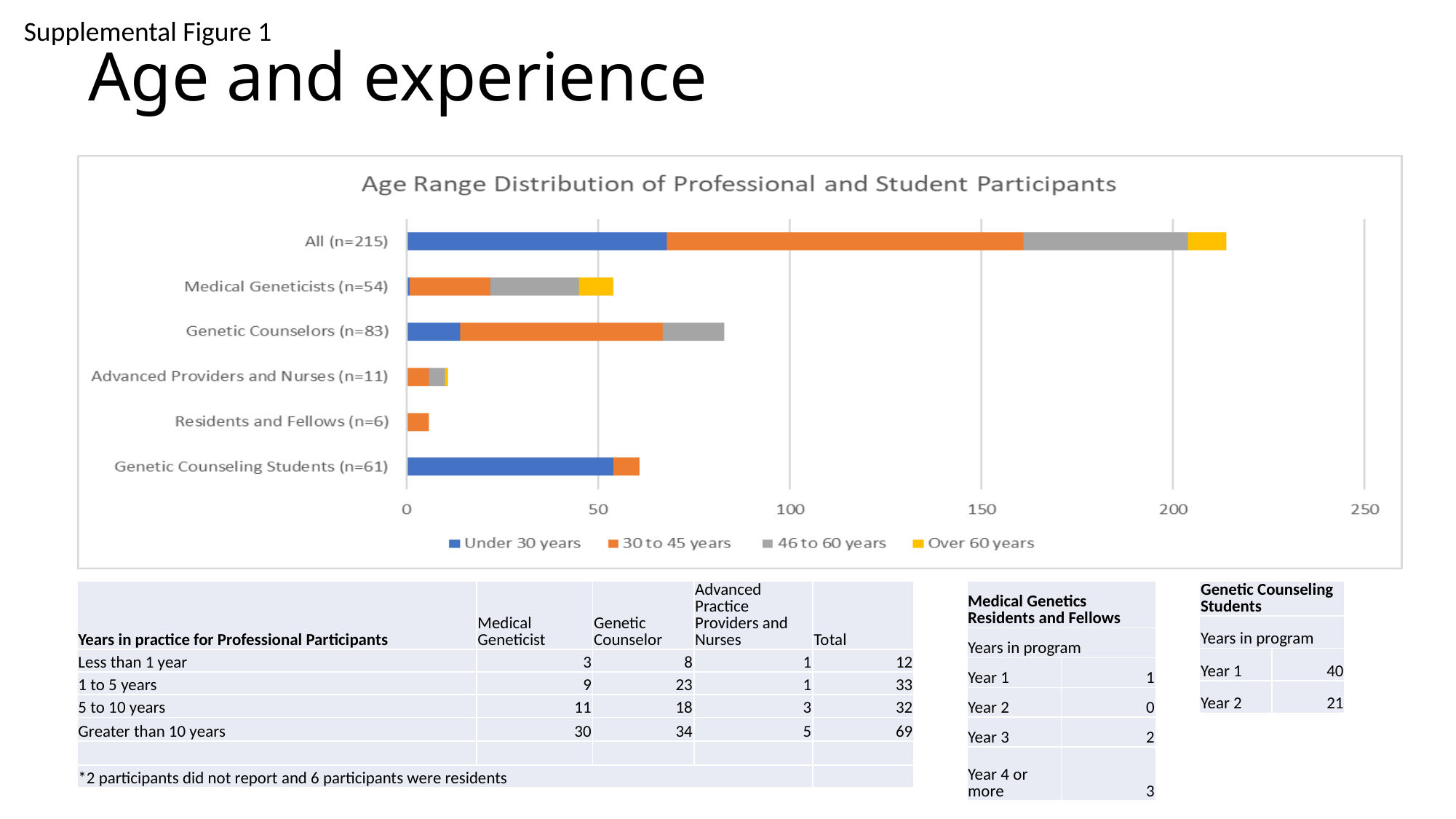

#### Age and experience
Supplemental Figure 1
| Years in practice for Professional Participants | Medical Geneticist | Genetic Counselor | Advanced Practice Providers and Nurses | Total |
| --- | --- | --- | --- | --- |
| Less than 1 year | 3 | 8 | 1 | 12 |
| 1 to 5 years | 9 | 23 | 1 | 33 |
| 5 to 10 years | 11 | 18 | 3 | 32 |
| Greater than 10 years | 30 | 34 | 5 | 69 |
| \*2 participants did not report and 6 participants were residents | | | | |
| Medical Genetics Residents and Fellows | |
| --- | --- |
| Years in program | |
| Year 1 | 1 |
| Year 2 | 0 |
| Year 3 | 2 |
| Year 4 or more | 3 |
| Genetic Counseling Students | |
| --- | --- |
| Years in program | |
| Year 1 | 40 |
| Year 2 | 21 |

### Slide 2
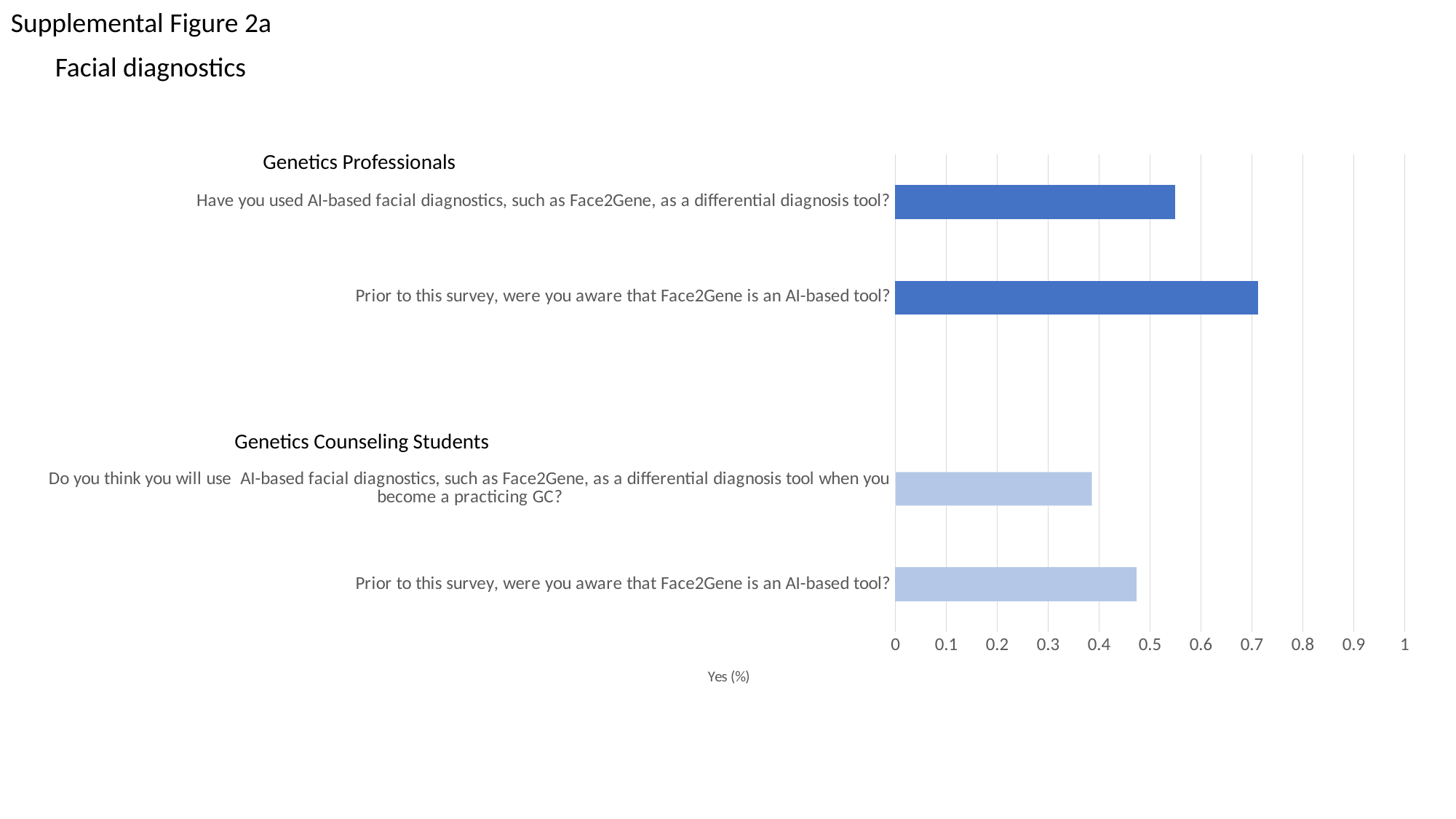

Supplemental Figure 2a
Facial diagnostics
###### Chart
| Category | |
|---|---|
| Prior to this survey, were you aware that Face2Gene is an AI-based tool? | 0.47368421052631576 |
| Do you think you will use AI-based facial diagnostics, such as Face2Gene, as a differential diagnosis tool when you become a practicing GC? | 0.38596491228070173 |
| | None |
| Prior to this survey, were you aware that Face2Gene is an AI-based tool? | 0.7124183006535948 |
| Have you used AI-based facial diagnostics, such as Face2Gene, as a differential diagnosis tool? | 0.5490196078431373 |Genetics Professionals
Genetics Counseling Students

### Slide 3
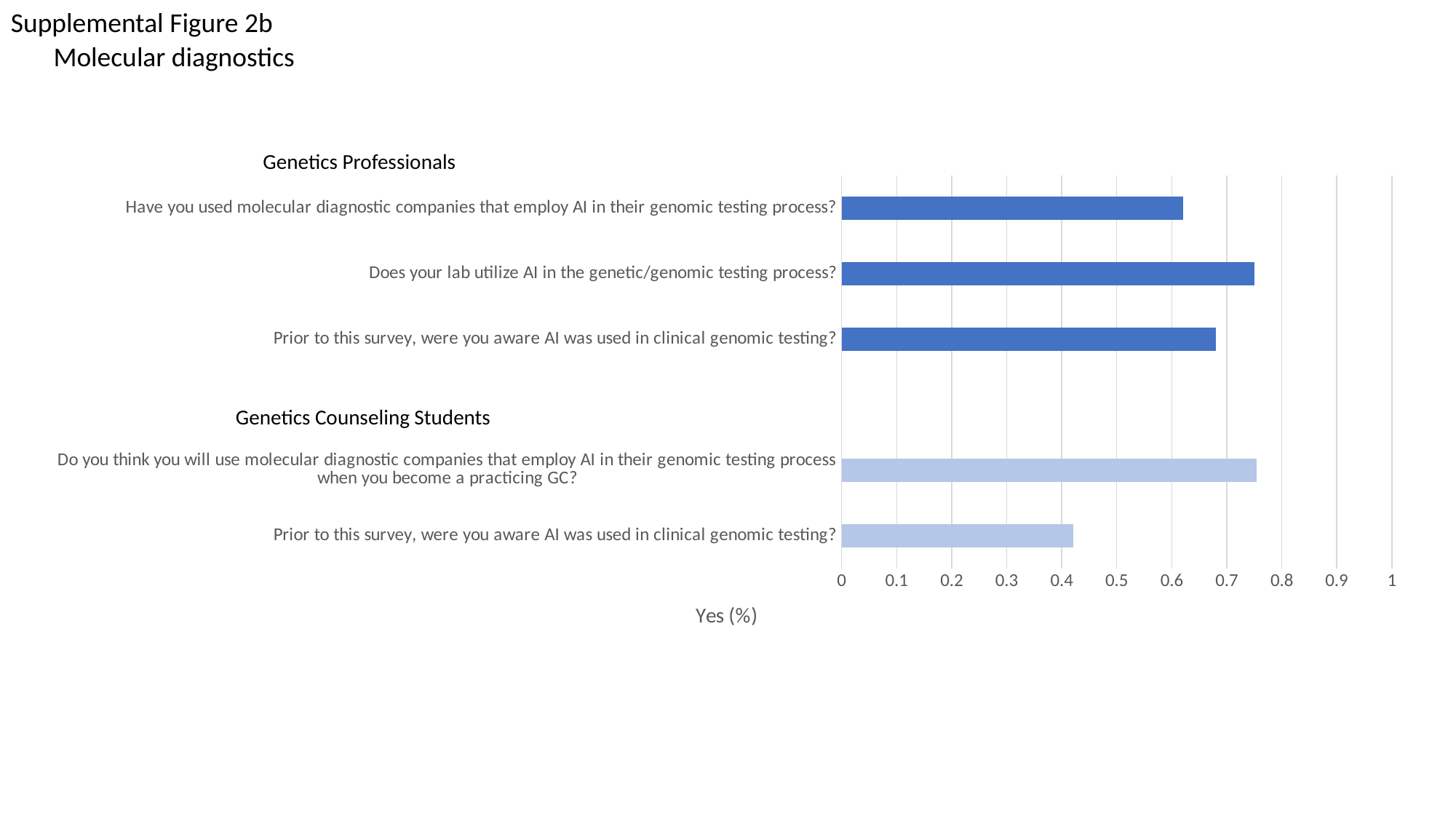

Supplemental Figure 2b
Molecular diagnostics
Genetics Professionals
###### Chart
| Category | |
|---|---|
| Prior to this survey, were you aware AI was used in clinical genomic testing? | 0.42105263157894735 |
| Do you think you will use molecular diagnostic companies that employ AI in their genomic testing process when you become a practicing GC? | 0.7543859649122807 |
| | None |
| Prior to this survey, were you aware AI was used in clinical genomic testing? | 0.6797385620915033 |
| Does your lab utilize AI in the genetic/genomic testing process? | 0.75 |
| Have you used molecular diagnostic companies that employ AI in their genomic testing process? | 0.6206896551724138 |Genetics Counseling Students

### Slide 4
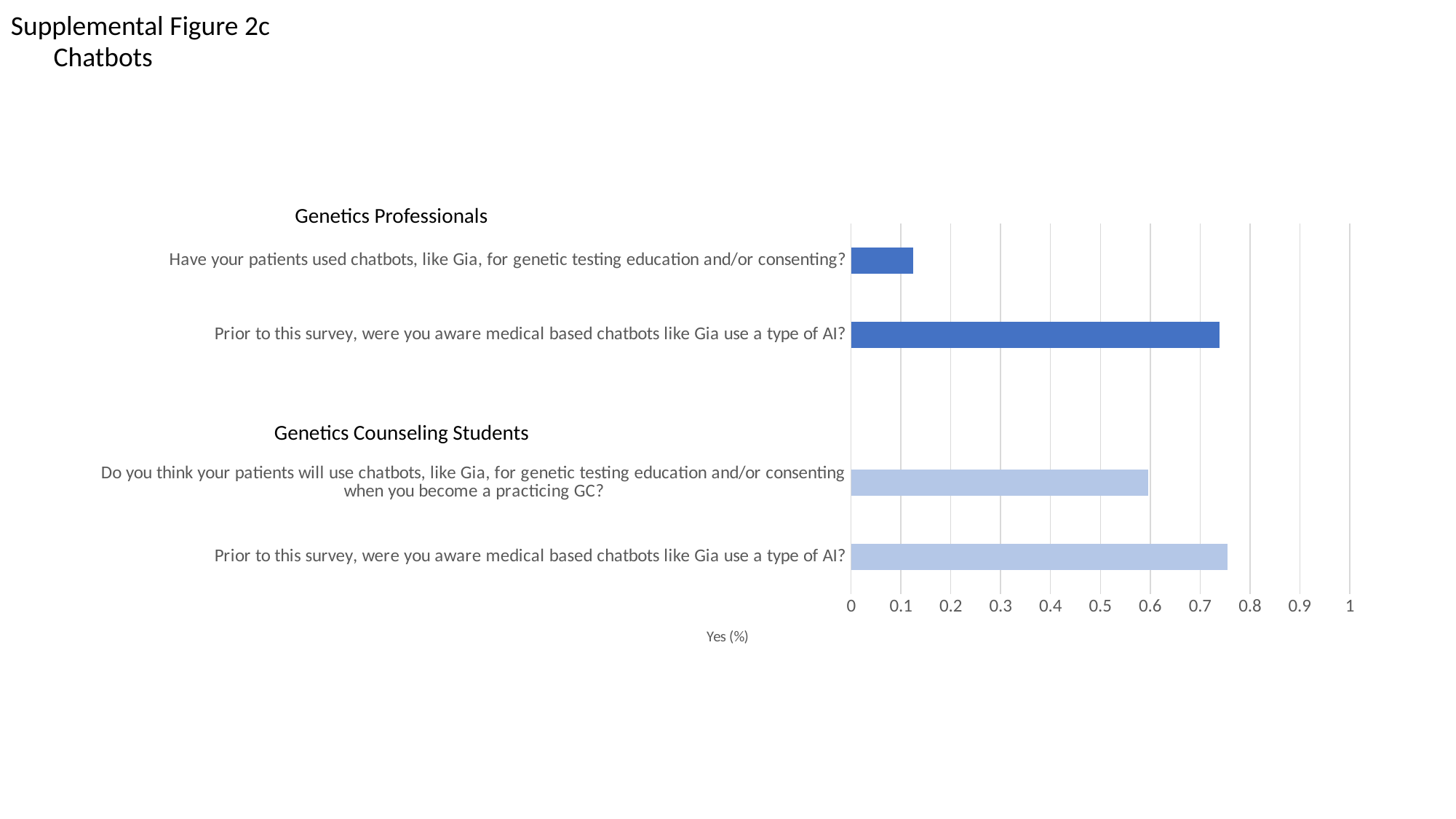

Supplemental Figure 2c
Chatbots
Genetics Professionals
###### Chart
| Category | |
|---|---|
| Prior to this survey, were you aware medical based chatbots like Gia use a type of AI? | 0.7543859649122807 |
| Do you think your patients will use chatbots, like Gia, for genetic testing education and/or consenting when you become a practicing GC? | 0.5964912280701754 |
| | None |
| Prior to this survey, were you aware medical based chatbots like Gia use a type of AI? | 0.738562091503268 |
| Have your patients used chatbots, like Gia, for genetic testing education and/or consenting? | 0.12418300653594772 |Genetics Counseling Students

### Slide 5
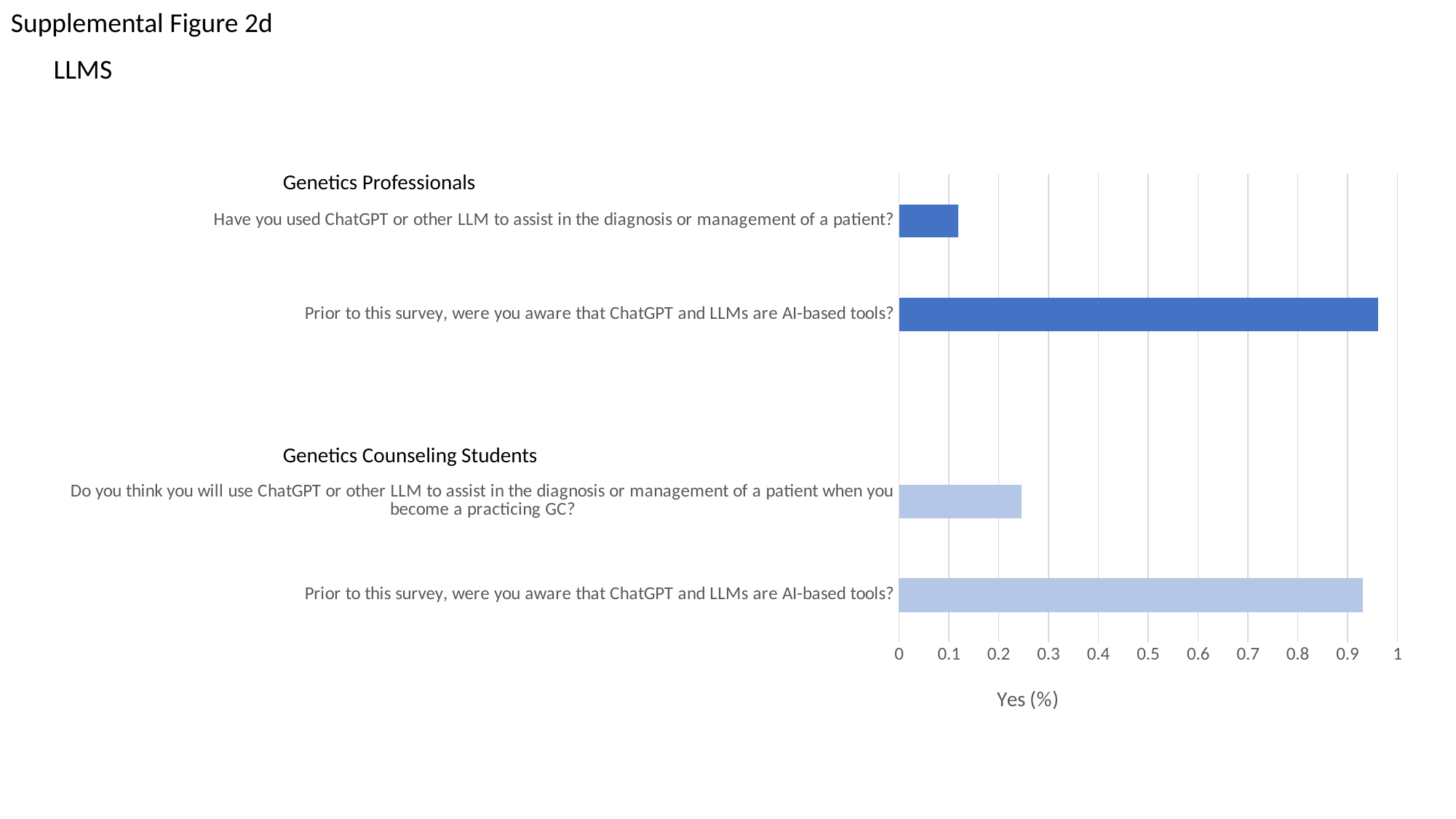

Supplemental Figure 2d
LLMS
###### Chart
| Category | |
|---|---|
| Prior to this survey, were you aware that ChatGPT and LLMs are AI-based tools? | 0.9298245614035088 |
| Do you think you will use ChatGPT or other LLM to assist in the diagnosis or management of a patient when you become a practicing GC? | 0.24561403508771928 |
| | None |
| Prior to this survey, were you aware that ChatGPT and LLMs are AI-based tools? | 0.9607843137254902 |
| Have you used ChatGPT or other LLM to assist in the diagnosis or management of a patient? | 0.11842105263157894 |Genetics Professionals
Genetics Counseling Students

### Slide 6
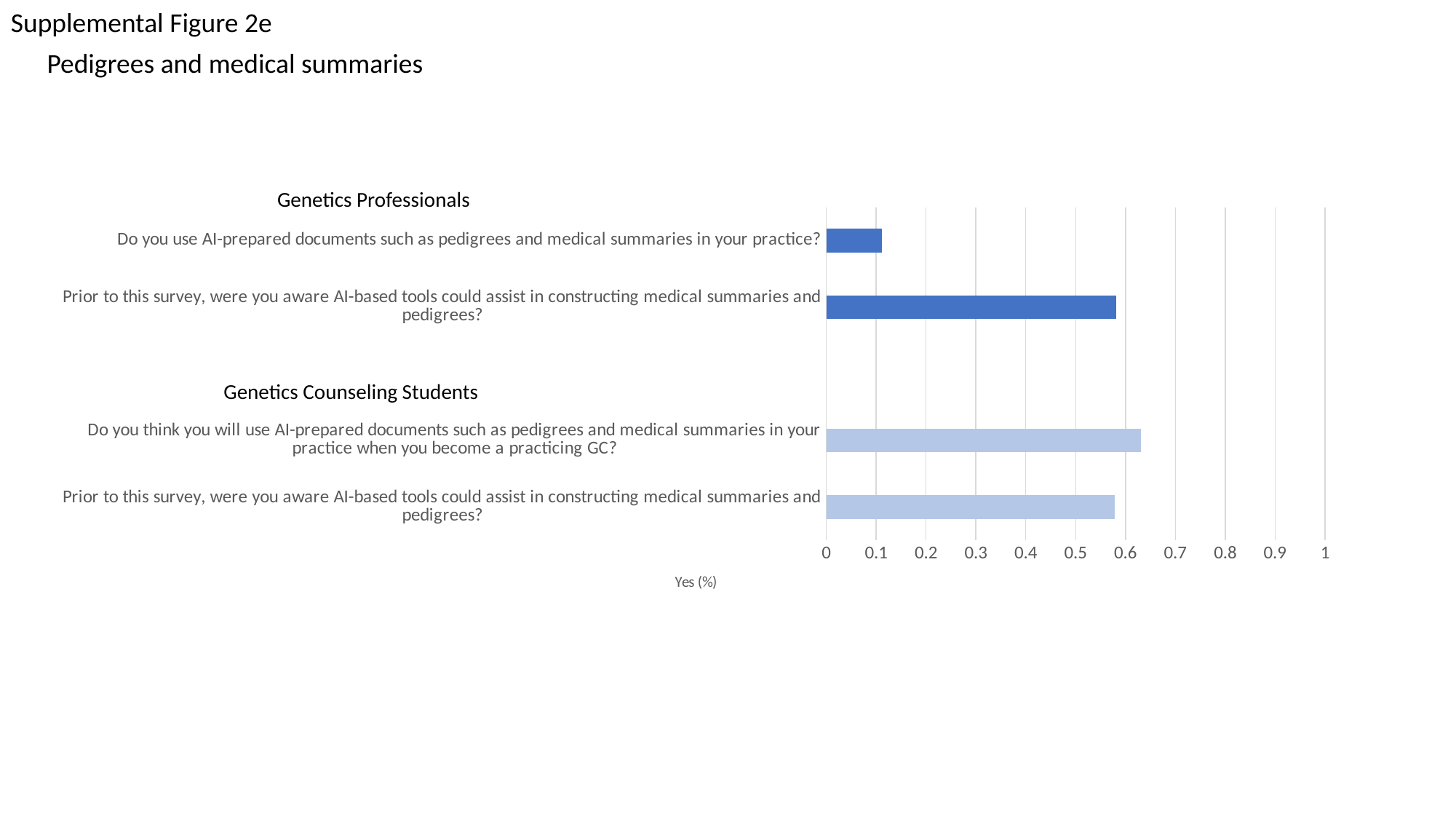

Supplemental Figure 2e
Pedigrees and medical summaries
Genetics Professionals
###### Chart
| Category | |
|---|---|
| Prior to this survey, were you aware AI-based tools could assist in constructing medical summaries and pedigrees? | 0.5789473684210527 |
| Do you think you will use AI-prepared documents such as pedigrees and medical summaries in your practice when you become a practicing GC? | 0.631578947368421 |
| | None |
| Prior to this survey, were you aware AI-based tools could assist in constructing medical summaries and pedigrees? | 0.5816993464052288 |
| Do you use AI-prepared documents such as pedigrees and medical summaries in your practice? | 0.1111111111111111 |Genetics Counseling Students

### Slide 7
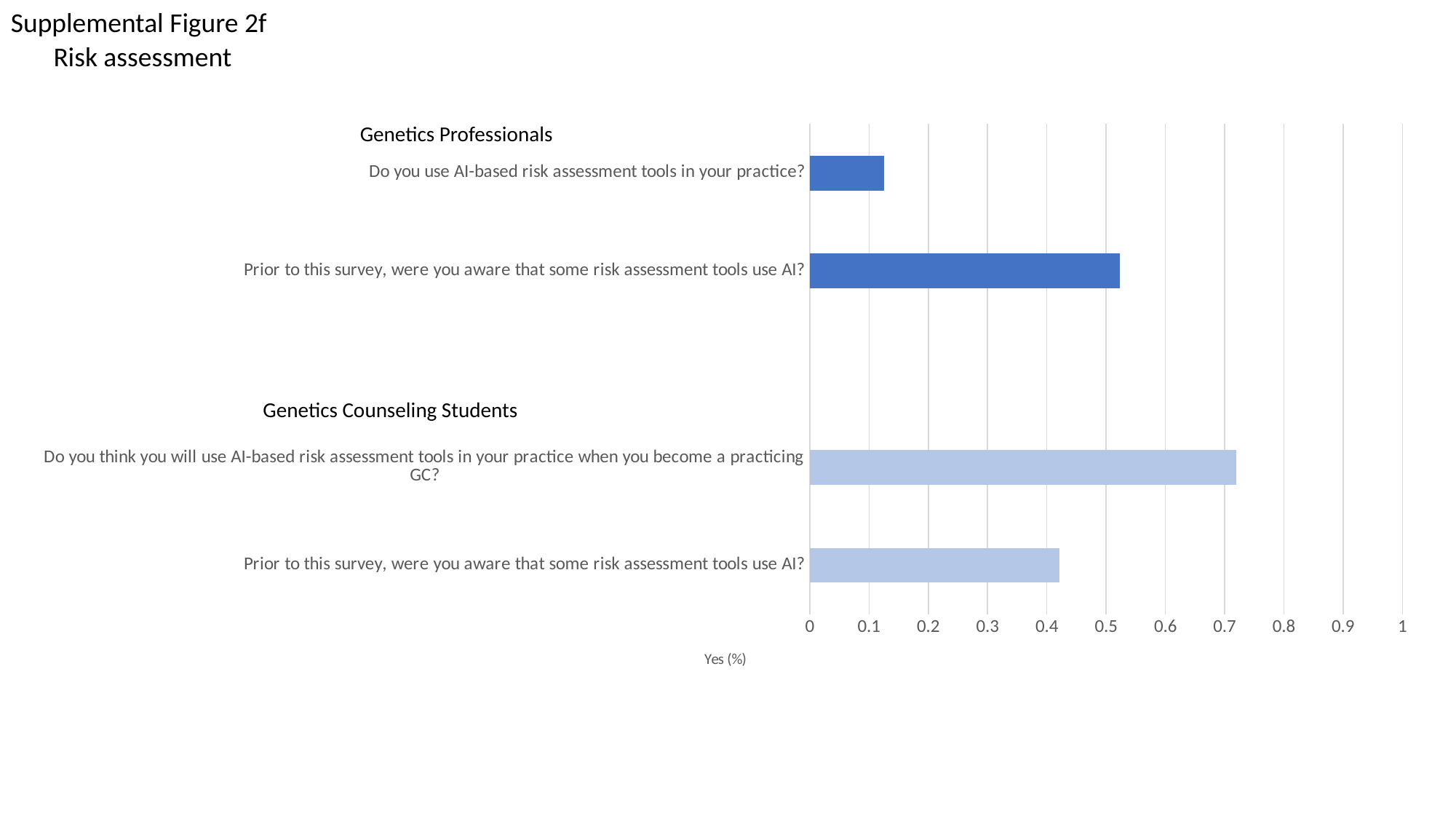

Supplemental Figure 2f
Risk assessment
###### Chart
| Category | |
|---|---|
| Prior to this survey, were you aware that some risk assessment tools use AI? | 0.42105263157894735 |
| Do you think you will use AI-based risk assessment tools in your practice when you become a practicing GC? | 0.7192982456140351 |
| | None |
| Prior to this survey, were you aware that some risk assessment tools use AI? | 0.5228758169934641 |
| Do you use AI-based risk assessment tools in your practice? | 0.125 |Genetics Professionals
Genetics Counseling Students

### Slide 8
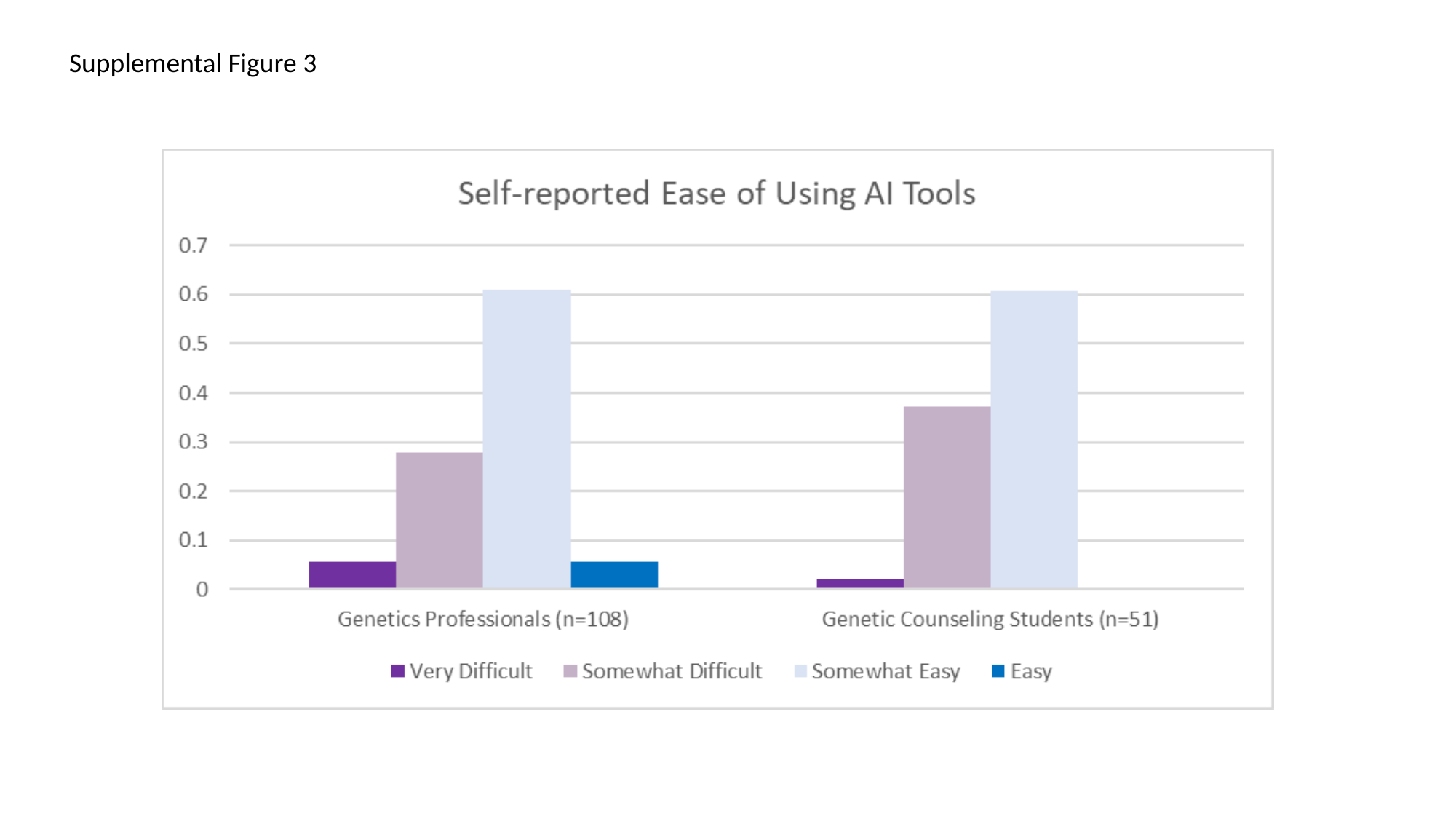

Supplemental Figure 3
