## Supplemental Table 1 for "Artificial intelligence in clinical genetics: current practice and attitudes among the clinical genetics workforce"

| Likert Scale Questions | | **Agree/**  **Strongly Agree** | **Disagree/ Strong Disagree** | **p-value** |
| --- | --- | --- | --- | --- |
| **Comfort with AI in Clinical Practice** | | | | |
| I am comfortable using AI in my clinical genetics practice. (%) | Clinicians n=150 | 94 (62.7) | 56 (37.3) | 0.87 |
|  | Students n=57 | 35 (61.4) | 22 (38.6) |  |
| My patients would be comfortable with the use of AI in clinical genetics. (%) | Clinicians n=147 | 97 (66) | 50 (34) | 4.4x10^-5^ |
|  | Students  n=56 | 19 (33.9) | 37 (66.1) |  |
| I am comfortable with my patients using AI to obtain general genetics information. (%) | Clinicians n=149 | 107 (71.8) | 42 (28.2) | 2.8x10^-3^ |
|  | Students n=56 | 27 (48.2) | 29 (51.8) |  |
| I am comfortable with my patients using AI to undergo pretest genetic counseling. (%) | Clinicians n=149 | 73 (49) | 76 (51) | 3.7x10^-4^ |
|  | Students n=56 | 13 (23.2) | 43 (76.8) |  |
| **AI Safety and Reliability** | | | | |
| AI provides reliable outputs. (%) | Clinicians n=146 | 105 (71.9) | 41 (28.1) | 0.63 |
|  | Students n=57 | 39 (68.4) | 18 (31.6) |  |
| I am concerned AI will not be used responsibly in clinical genetics. (%) | Clinicians n=149 | 66 (44.3) | 83 (55.7) | 9.3x10^-3^ |
|  | Students n=57 | 36 (63.2) | 21 (36.8) |  |
| There needs to be further regulations for AI before I incorporate it into my genetics practice. (%) | Clinicians n=148 | 113 (76.4) | 35 (23.6) | 4.8x10^-6^ |
|  | Students n=57 | 55 (96.5) | 2 (3.5) |  |
| AI can be safely used in the handling of confidential data. (%) | Clinicians n=147 | 101 (68.7) | 46 (31.3) | 0.23 |
|  | Students n=57 | 29 (50.9) | 28 (49.1) |  |
| **Clinical Genetics Clinic Workload** | | | | |
| Clinical genetics professionals are over-burdened. (%) | Clinicians n=147 | 138 (93.9) | 9 (6.1) | 0.41 |
|  | Students n=57 | 55 (96.5) | 2 (3.5) |  |
| I am willing to delegate work to other people. (%) | Clinicians n=147 | 138 (93.9) | 9 (6.1) | 0.78 |
|  | Students n=55 | 51 (92.7) | 4 (7.3) |  |
| I am willing to delegate work to AI. (%) | Clinicians n=146 | 111 (76) | 35 (24) | 1.1x10^-3^ |
|  | Students n=55 | 51 (92.7) | 4 (7.3) |  |
| AI is an appropriate solution to relieving workload. (%) | Clinicians n=146 | 108 (74) | 38 (26) | 0.55 |
|  | Students n=56 | 39 (69.6) | 17 (30.4) |  |
| **Attitudes Toward AI Tools** | | | | |
| I have felt forced to utilize AI in my practice. (%) | Clinicians n=150 | 11 (7.3) | 139 (92.7) | 0.63 |
|  | Students n=57 | 4 (7) | 53 (93) |  |
| Using AI as an aid is outside my scope of training/practice. (%) | Clinicians n=149 | 38 (25.5) | 111 (74.5) | 0.26 |
|  | Students n=57 | 19 (33.3) | 38 (66.7) |  |
| AI is not ready to be used in clinical genetics. (%) | Clinicians n=147 | 31 (21.1) | 116 (78.9) | 0.02 |
|  | Students n=57 | 21 (36.8) | 36 (63.2) |  |
| AI is inappropriate for use as a diagnostic support tool. (%) | Clinicians n=151 | 21 (13.9) | 130 (86.1) | 0.23 |
|  | Students n=57 | 12 (21.1) | 45 (78.9) |  |
| The development of new medical genetics AI tools requires input from clinical genetics professionals. (%) | Clinicians n=149 | 149 (100) | 0 | 0.32 |
|  | Students n=57 | 56 (98.2) | 1 (1.8) |  |
| **AI Knowledge and Education** | | | | |
| I currently lack sufficient knowledge about AI in medical genetics. (%) | Clinicians n=149 | 116 (78.4) | 32 (21.6) | 2.4x10^-5^ |
|  | Students n=57 | 55 (96.5) | 2 (3.5) |  |
| There should be further curriculum and education about the use of AI in clinical care. (%) | Clinicians  n=149 | 146 (98) | 3 (2) | 0.53 |
|  | Students n=57 | 55 (96.5) | 2 (3.5) |  |
| If offered a class about AI in clinical genetics, I would take it. (%) | Clinicians n=148 | 129 (86.5) | 20 (13.5) | 0.04 |
|  | Students n=57 | 55 (96.5) | 2 (3.5) |  |

Supplemental Table 1. Comparison of clinicians and students for Likert scale question.
