## Supplemental Table 2 for "Artificial intelligence in clinical genetics: current practice and attitudes among the clinical genetics workforce"

|  | Medical Geneticists | Genetic Counselors | Physician Assistant | Medical Genetics Residents/Fellows | Genetic Counseling Students |
| --- | --- | --- | --- | --- | --- |
| Received AI training in clinical program (%) | 2/53  (3.8) | 4/82  (4.9) | 1/9  (11.1) | 1/6  (16.7) | 12/61  (19.7) |
| Received AI training in workshop/seminar (%) | 16/54  (29.6) | 28/83  (33.7) | 2/9  (22.2) | 1/6  (16.7) | 20/61  (32.8) |
| Rated AI clinical knowledge as intermediate/advanced (%) | 37/55  (66.7) | 43/83  (51.8) | 5/9  (55.6) | 2/6  (33.3%) | 19/59  (32.2) |
| AI tools are somewhat easy to very easy to use. (%) | 29/48  (60.4) | 47/68  (69.1) | 3/6  (50) | 5/6  (83.3) | 35/56  (62.5) |
| Use AI in clinical practice (before tutorial) (%) | 29/54  (53.7) | 36/83  (43.4) | 5/9  (55.6) | 4/6  (66.7) | 49/61*  (80.3) |
| Clinical AI tools accessible/ most tools accessible (before tutorial) (%) | 31/54  (57.4) | 27/84  (32.1) | 3/9  (33.3) | 4/6  (66.7) | 20/61  (32.8) |
| Clinical AI tools beneficial/highly beneficial (before tutorial) (%) | 44/50  (88) | 65/76  (85.5) | 8/9  (88.9) | 5/6  (83.3) | 48/60  (80) |

Supplemental Table 2. Responses according to profession or training program. Results for the one registered nurse and one nurse practitioner not included in this table.

*Student were not asked about current use but instead asked about anticipated use once a practicing clinician.
