## Supplemental Table 3-4 for "Artificial intelligence in clinical genetics: current practice and attitudes among the clinical genetics workforce"

|  | Medical Geneticists | Genetic Counselors | Physician Assistant | Medical Genetics Residents/Fellows | Genetic Counseling Students |
| --- | --- | --- | --- | --- | --- |
| Likert Scale Questions | Agree/Strongly Agree with the Likert Question Statement | | | | |
| I am comfortable using AI in my clinical genetics practice. (%) | 32/53  (60.4) | 52/80  (65) | 5/9  (55.6) | 3/6  (50) | 35/57 (61.4) |
| My patients would be comfortable with the use of AI in clinical genetics. (%) | 36/53  (67.9) | 52/77  (67.5) | 5/9  (55.6) | 3/6  (50) | 19/56 (33.9) |
| I am comfortable with my patients using AI to obtain general genetics information. (%) | 34/53  (64.2) | 64/79  (81) | 5/9  (55.6) | 3/6  (50) | 27/56 (48.2) |
| I am comfortable with my patients using AI to undergo pretest genetic counseling. (%) | 27/53  (50.9) | 38/79  (48.1) | 3/9  (33.3) | 4/6  (66.7) | 13/56 (23.2) |
| AI provides reliable outputs. (%) | 27/52  (51.9) | 66/78  (84.6) | 5/8  (62.5) | 5/6  (83.3) | 39/57 (68.4) |
| I am concerned AI will not be used responsibly in clinical genetics. (%) | 23/52  (44.2) | 37/80  (46.3) | 2/9  (22.2) | 3/6  (50) | 36/57 (63.2) |
| There needs to be further regulations for AI before I incorporate it into my genetics practice. (%) | 36/53  (67.9) | 77/79  (97.5) | 9/9  (100) | 4/6  (66.7) | 55/57 (96.5) |
| AI can be safely used in the handling of confidential data. (%) | 33/52  (63.5) | 56/78  (71.8) | 6/9  (66.7) | 4/6  (66.7) | 2/579 (50.9) |
| Clinical genetics professionals are over-burdened. (%) | 48/52  (92.3) | 77/79  (97.5) | 6/8  (75) | 5/6  (83.3) | 55/57 (96.5) |
| I am willing to delegate work to other people. (%) | 48/53  (90.6) | 74/44  (96.1) | 9/9  (100) | 5/6  (83.3) | 51/55  (92.7) |
| I am willing to delegate work to AI. (%) | 39/53  (73.6) | 58/76  (76.3) | 7/9  (77.8) | 6/6  (100) | 51/55  (92.7) |
| AI is an appropriate solution to relieving workload. (%) | 36/53  (67.9) | 59/77  (76.6) | 6/8  (75) | 5/6  (83.3) | 39/56 (69.6) |
| I have felt forced to utilize AI in my practice. (%) | 4/53  (7.5) | 6/80  (7.5) | 0/9  (0) | 1/6  (16.7) | 4/57  (7) |
| Using AI as an aid is outside my scope of training/practice. (%) | 17/53  (32.1) | 17/79  (21.5) | 1/9  (11.1) | 1/6  (16.7) | 19/57 (33.3) |
| AI is not ready to be used in clinical genetics. (%) | 12/52  (23.1) | 16/78  (20.5) | 2/9  (22.2) | 1/6  (16.7) | 21/57 (36.8) |
| AI is inappropriate for use as a diagnostic support tool. (%) | 10/53  (18.9) | 10/80  (12.5) | 1/9  (11.1) | 0/6  (0) | 12/57 (21.1) |
| The development of new medical genetics AI tools requires input from clinical genetics professionals. (%) | 53/53  (100) | 79/79  (100) | 9/9  (100) | 6/6  (100) | 56/57 (98.2) |
| I currently lack sufficient knowledge about AI in medical genetics. (%) | 36/53  (67.9) | 67/78  (85.9) | 6/9  (66.7) | 5/6  (83.3) | 55/57 (96.5) |
| There should be further curriculum and education about the use of AI in clinical care. (%) | 52/53  (98.1) | 77/79  (97.5) | 9/9  (100) | 6/6  (100) | 55/57 (96.5) |
| If offered a class about AI in clinical genetics, I would take it. (%) | 44/53  (83) | 71/79  (89.9) | 8/9  (88.9) | 6/6  (100) | 55/57 (96.5) |

Supplemental Table 3. Agreement responses according to profession or training program. Results for the one registered nurse and one nurse practitioner not included in this table due to limited numbers of these participant groups.

|  | Comparison of Medical Geneticist and Genetic Counselor Responses  p-value |
| --- | --- |
| Clinical AI tools accessible/ most tools accessible (before tutorial) (%) | 0.005 |
| I am comfortable with my patients using AI to obtain general genetics information. – Agree with statement (%) | 0.03 |
| AI provides reliable outputs. – Agree with statement (%) | 3.42x10^-5^ |
| There needs to be further regulations for AI before I incorporate it into my genetics practice. – Agree with statement (%) | 2.05x10^-8^ |
| I currently lack sufficient knowledge about AI in medical genetics. – Agree with statement (%) | 0.01 |

Supplemental Table 4. Significant differences in medical geneticist and genetic counselor responses.
